# Comorbidities, geriatric syndromes, and glycaemic control among older patients with diabetes: a multi-centre study in Vietnam

**DOI:** 10.64898/2025.12.03.25341581

**Authors:** Wei Jin Wong, Tan Van Nguyen, Dung Ngoc Truong, Ying Zhang, Christopher Harrison, Mark Woodward, Tu Ngoc Nguyen

## Abstract

**Aim:** This study aimed to examine the burden of comorbidities and common geriatric syndromes in older adults with type 2 diabetes mellitus in Vietnam and their relationship with poor glycaemic control.

**Methods:** This is a cross-sectional study of patients aged 60 years or older diagnosed with type 2 diabetes who visited the cardio-metabolic clinics of two urban hospitals in Ho Chi Minh City, Vietnam, between November 2022 and June 2023. Poor glycaemic control was defined as HbA1c ≥ 7.0%. Comorbidity severity was assessed using the Charlson Comorbidity Index (CCI). Frailty was defined using Fried’s criteria. Polypharmacy was defined as the use of five or more medications daily. Multiple-adjusted logistic regression models were applied to identify the factors associated with poor glycaemic control.

**Results:** There were 576 participants. They had a mean age of 71.9 (SD 7.6) years, 46% were female, 30% were frail, 77% had polypharmacy. The mean duration of diabetes was 10.9 years (SD 7.4). The mean CCI was 2.5 (SD 1.1), and 34% of the participants had a CCI ≥3. The most common comorbidities were dyslipidaemia (98%), hypertension (96%), followed by chronic kidney disease (17%), and peripheral artery disease (11%). The prevalence of poor glycaemic control (HbA1c ≥ 7%) was 47%. The factors significantly associated with poor glycaemic control were polypharmacy, frailty, and long duration of diabetes.

**Conclusion:** There was a high prevalence of comorbidity, frailty, and polypharmacy among the participants; all were related to poor glycaemic control. Optimizing polypharmacy, frailty and comorbidity is essential in managing long-term diabetes.

## Introduction

Globally, the prevalence of diabetes among people aged 65 years and older is approximately 20%.^1^ It is anticipated that, by 2045, approximately 75% of adults with type 2 diabetes mellitus (T2DM) will be living in low- and middle-income countries (LMICs).^2,3^ Countries around the world are undergoing an epidemiological transition, as populations age. This trend is particularly pronounced in LMICs, where the growth rate of older populations is expected to surpass high-income countries by 2050.^4^ As diabetes can increase the risk of other chronic health conditions, such as cardiovascular and kidney diseases, it continues to be a significant public health challenge. Effective glycaemic control becomes essential for preventing acute complication and reducing risks of long-term micro- and macro-vascular outcomes. However, achieving and maintaining glycaemic control is a challenge due to a variety of influencing factors. Glycaemic control among individuals with T2DM in LMICs have previously been reported as suboptimal.^5^

Older adults often have complex health needs that can make their diabetes care more challenging. ^6 7^ For example, increased risk of hypoglycemia from commonly used medicines like metformin and sulfonylureas due to reduced kidney function that happens with aging. In severe stages of reduced kidney function, these medicines are sometimes contraindicated limiting management options for older adults. The current management of T2DM focuses mainly on controlling blood glucose levels assessed by haemoglobin A1c (HbA1c) levels. The American Diabetes Association Standards of Care in Diabetes 2025 ^8^ highlights the need for individualized goals in older adults due to the heterogeneity in health status, functional abilities, and cognitive function in this population. For the general adult population, it recommended HbA1c targets of < 6.5% when achievable without significant hypoglycaemia, or under 7.0% for most individuals at high risk of hypoglycaemia. For older adults with T2DM and intermediate or complex health (such as frailty, cognitive or functional impairments, severe coexisting conditions), a more relaxed target (HbA1c < 8.0%) was recommended. The International Geriatric Diabetes Society further highlights how care provides for older adults with diabetes should respond to changes associated with aging, such as increasing frailty, cognitive impairment and comorbidities which may affect glycaemic management.^6^

In Vietnam, the prevalence of T2DM has been steadily increasing.^9^ Recent reports indicate a rising prevalence of prediabetes and undiagnosed T2DM, with older age as one of the associated factors.^10^ Studies have also reported that the rate of poor glycaemic control in Vietnam ranged from 33% - 82%.^11^ Poor glycaemic control is more frequently observed in older adults with T2DM, who often experience multiple geriatric conditions, such as frailty, multimorbidity and polypharmacy.^12,13^ However, there is a lack of studies that focus on these issues in older adults with T2DM in Vietnam, and specifically how these geriatric conditions affect diabetes management in this population.

This study aimed to examine the burden of comorbidities and common geriatric syndromes (frailty, polypharmacy) in older adults with type 2 diabetes in Vietnam and their relationship with poor glycaemic control.

## Methods

### Study design and population

A cross-sectional analysis was conducted, using data from a cohort study of frailty in older patients with type 2 diabetes in Vietnam. Details of this study were published elsewhere. Briefly, a total of 644 older patients aged 60 years or older diagnosed with type 2 diabetes who visited the cardio-metabolic clinics of Thong Nhat Hospital and Gia Dinh Hospital in Ho Chi Minh City from November 2022 to June 2023 were recruited. Among these 644 participants, those with a measurement of glycated haemoglobin (HbA1c) (n=576) were included in the current study.

### Data collection

Data were collected from patient interviews and medical records. Information obtained included demographic characteristics, height, weight, medical history, duration of having diabetes (in years), number of medications used, and number of comorbidities. Body mass index (BMI) was calculated from measured weight and height and classified into four groups: underweight (BMI < 18.5 kg/m^2^), ideal (BMI 18.5–22.9 kg/m^2^), overweight (BMI 23.0–24.9 kg/m^2^), and obese (BMI ≥ 25.0 kg/m^2^). Comorbidities were assessed using the Charlson Comorbidity Index (CCI), which estimates the 10-year risk of death associated with a combination of comorbidities. Comorbidity strength was categorized as mild (CCI 1-2), moderate (CCI 3-4), and severe (CCI ≥ 5).^14^ Frailty was defined by Fried’s frailty criteria, which include five components (unintentional weight loss, weakness, exhaustion, slowness and low physical activity).^15^ Participants with three or more of these five components were identified as being frail, those with one or two components as prefrail, and those with none as robust. Polypharmacy was defined as the use of five or more medications daily.^16^

HbA1c values were obtained from the latest measurement recorded within the past three months in the medical records. Poor glycaemic control was defined as HbA1c ≥7.0%.^8^

The studies were approved by the Ethics Committee of the University of Medicine and Pharmacy at Ho Chi Minh City (Reference Number 934/DHYD-HDDD, dated 24/11/2022). Informed consent was obtained from all participants.

### Statistical analysis

Participant characteristics are presented as mean and standard deviation (SD) for continuous variables, and frequencies and percentages for categorical variables. Comparisons among groups were conducted using chi-square tests or Fisher’s exact tests for binary variables, and Student’s t-tests or One-Way ANOVA for continuous variables.

Logistic regression models were applied to identify the factors associated with poor glycaemic control. Unadjusted odds ratios (OR) and adjusted ORs and their 95% confidence intervals (CIs) were calculated for the predefined variables. Besides three main variables of interest (frailty, polypharmacy, and the Charlson Comorbidity Index), we also included socio-demographic factors (age, sex, marital status, educational level), lifestyle factor (smoking), duration of diabetes, and cardiovascular risk factors/cardiovascular disease (hypertension, dyslipidaemia, obesity, heart failure, coronary heart disease, stroke, peripheral artery disease, chronic kidney disease). P values <0.05 were considered statistically significant. All variables were examined for interaction and multicollinearity. Data were analysed in SPSS Statistics 29.0.

## Results

### Characteristics of the participants

The 576 participants had a mean age of 71.9 (SD 7.6) years, 46% were female, 96% were retired, and 74% were married. Regarding educational level, 9% had low education (completed primary school or less), 47% completed high school, 36% attended college or university, and 9% obtained higher education. In terms of diabetes duration, 36% of the participants had diabetes for more than 10 years, followed by 6-10 years (32%), 1-5 years (28%) and < 1 year (4%).

### Prevalence of comorbidities, polypharmacy, and frailty

The most common cardiovascular comorbidities were dyslipidaemia (98%), hypertension (96%), chronic kidney disease (17%), peripheral artery disease (11%), coronary heart disease (2%), ischemic stroke (2%) and heart failure (1%), with no significant differences among participants with good and poor glycaemic control (Table 1).

**Table 1.** Participant characteristics.

| **Characteristic** | **All participants**<br>**(N = 576)** | **Participants**<br>**with HbA1c <**<br>**7%**<br>**(n=306)** | **Participants**<br>**with HbA1c ≥**<br>**7%**<br>**(n=270)** | **p-value** |
| --- | --- | --- | --- | --- |
| Age, years | 71.9 (7.6) | 72.1 (7.4) | 71.7 (7.9) | 0.232 |
| Age group |  |  |  |  |
| < 75 | 383 (67) | 204 (67) | 179 (66) | 0.925 |
| ≥ 75 | 193 (34) | 102 (33) | 91 (34) |  |
| Sex |  |  |  |  |
| Male | 309 (54) | 165 (54) | 144 (53) | 0.888 |
| Female | 267 (46) | 141 (46) | 126 (47) |  |
| Working status |  |  |  |  |
| Working | 24 (4) | 13 (4) | 11 (4) | 0.917 |
| Retired | 552 (96) | 293 (96) | 259 (96) |  |
| Marital status |  |  |  |  |
| Married | 426 (74) | 226 (74) | 200 (74) | 0.679 |
| Never married | 16 (3) | 7 (2) | 9 (3) |  |
| Divorced/separated | 7 (1) | 5 (2) | 2 (1) |  |
| Widowed | 127 (22) | 68 (22) | 59 (22) |  |
| Education |  |  |  |  |
| Primary school or less | 49 (9) | 26 (9) | 23 (9) | 0.056 |
| High school | 269 (47) | 158 (52) | 111 (41) |  |
| College/University | 206 (36) | 100 (33) | 106 (39) |  |
| Higher education | 52 (9) | 22 (7) | 30 (11) |  |
| Body mass index |  |  |  |  |
| Underweight | 15 (3) | 10 (3) | 5 (2) | 0.722 |
| Normal | 309 (54) | 160 (53) | 149 (55) |  |
| Overweight | 153 (27) | 82 (27) | 71 (26) |  |
| Obese | 96 (17) | 51 (17) | 45 (17) |  |
| Smoking (yes vs. no) | 166 (29) | 88 (29) | 78 (29) | 0.972 |
| Duration of diabetes<br>(years) | 10.9 (7.4) | 9.7 (7.1) | 12.3 (7.6) | <0.001 |
| <1 year | 23 (4) | 12 (4) | 11 (4) | <0.001 |
| 1 – 5 years | 161 (28) | 107 (35) | 54 (20) |  |
| 6 – 10 years | 186 (32) | 96 (31) | 90 (33) |  |
| >10 years | 206 (36) | 91 (30) | 115 (43) |  |
| Total number of medications | 5.5 (1.4) | 5.3 (1.4) | 5.8 (1.3) | <0.001 |
| Polypharmacy | 441 (77) | 215 (70) | 226 (84) | <0.001 |
| Frailty |  |  |  |  |
| Robust | 141 (25) | 90 (29) | 51 (19) | 0.011 |
| Pre-frail | 265 (46) | 135 (44) | 130 (48) |  |
| Frail | 170 (30) | 81 (27) | 89 (33) |  |
| Charlson Comorbidity Index (CCI) | 2.5 (1.1) | 2.4 (1.2) | 2.5 (1.1) | 0.463 |
| Comorbidity severity |  |  |  |  |
| CCI 1 – 2 (mild) | 379 (66) | 204 (67) | 175 (65) | 0.715 |
| CCI 3 – 4 (moderate) | 160 (28) | 81 (27) | 79 (29) |  |
| CCI ≥ 5 (severe) | 37 (6) | 21 (7) | 16 (6) |  |
| Cardiovascular risk factors and comorbidities |  |  |  |  |
| Dyslipidaemia | 564 (98) | 302 (99) | 262 (97) | 0.242 |
| Hypertension | 551 (96) | 291 (95) | 260 (96) | 0.481 |
| Chronic kidney disease | 96 (17) | 50 (16) | 46 (17) | 0.823 |
| Peripheral artery disease | 65 (11) | 34 (11) | 31 (12) | 0.889 |
| Coronary heart disease | 10 (2) | 7 (2) | 3 (1) | 0.349 |
| Ischemic stroke | 9 (2) | 7 (2) | 2 (1) | 0.135 |
| Heart failure | 8 (1) | 5 (2) | 3 (1) | 0.729 |

Continuous data are presented as mean (standard deviation). Categorical data are shown as n (%). CCI: Charlson Comorbidity Index, HbA1c: Glycated hemoglobin

Regarding the overall comorbidity burden, more than a third (34%) of all participants had moderate to severe comorbidity (28% reporting CCI of 3-4 and 6% reporting a CCI of ≥ 5), while 66% reporting a CCI of 1-2, and there was no significant differences among participants with good and poor glycaemic control.

The prevalence of polypharmacy was 77% in all participants, 70% in those with good glycaemic control vs. 84% in those with poor glycaemic control (p<0.001).

The prevalence of frailty and pre-frailty was 30% and 46% in all participants; 27% and 44% in those with good glycaemic control vs. 33% and 48% in those with poor glycaemic control, respectively (p=0.011)

### Mean levels of HbA1c by duration of diabetes, the severity of comorbidities, polypharmacy, and frailty

The prevalence of poor glycaemic control (HbA1c ≥ 7%) was 47%.

The distribution of HbA1c levels among the study participants is presented in Figure 1. Overall, the mean HbA1c level was 7.3%, SD 1.5.

**Figure 1.**
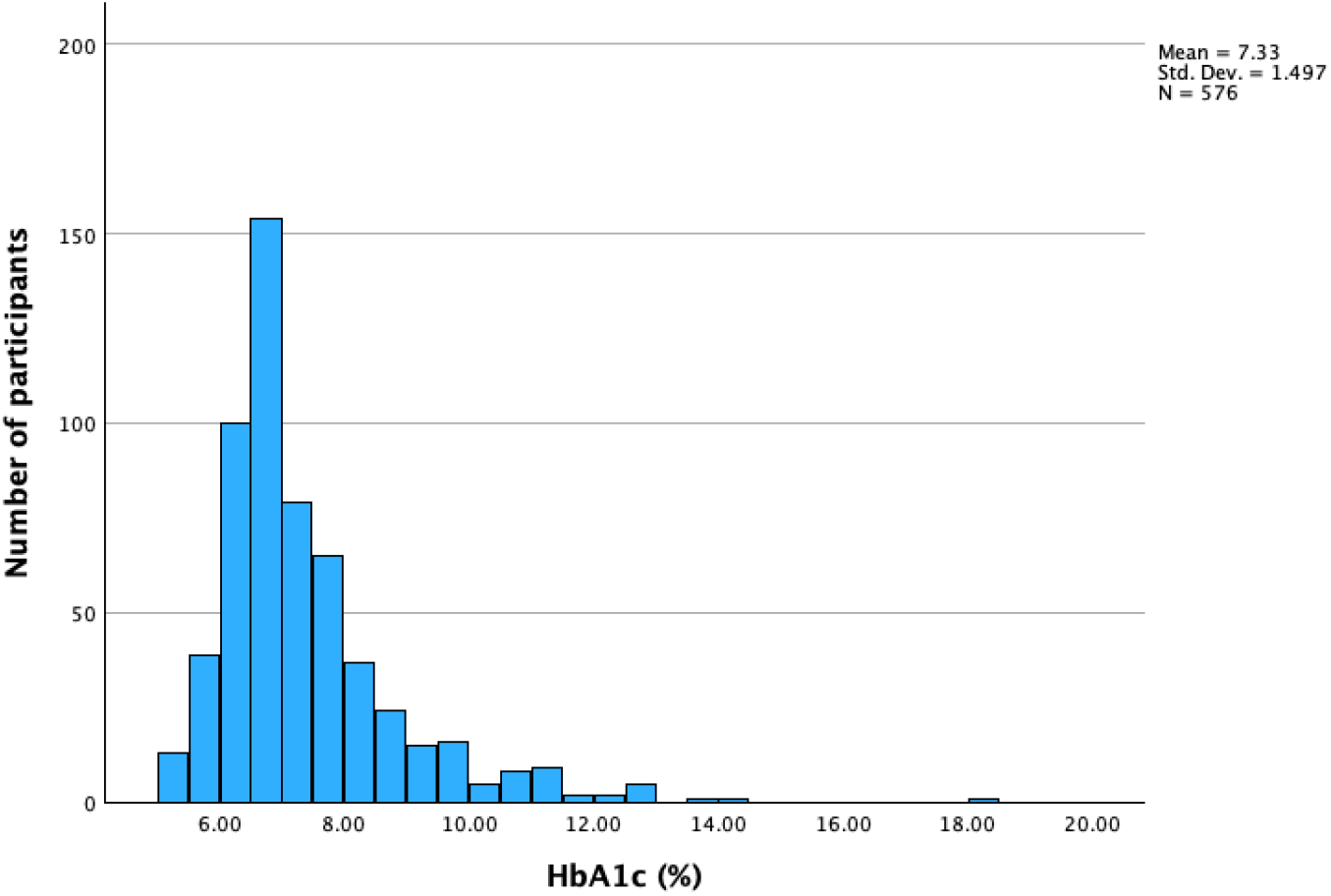
The distribution of HbA1c among the study participants

HbA1c level was highest among participants that were diagnosed with diabetes for less than 1 year (7.8%, SD 1.9), followed by those with more than 10 years of diabetes (7.5%, SD 1.4), 6-10 years (7.4%, SD 1.6), and lowest among those with 1-5 years of diabetes (7.0%, SD 1.4), p=0.004.

The mean HbA1c level was significantly higher in participants with polypharmacy compared to those without polypharmacy (7.4%, SD 1.4 vs. 7.1%, SD 1.6, p=0.018).

There were no significant differences by frailty (7.1%, SD 1.2 in robust, 7.3%, SD 1.6 in prefrail, and 7.5%, SD 1.5 in frail participants, p=0.074), and by comorbidity severity (7.3%, SD 1.4 in those with CCI 1-2, 7.5%, SD 1.7 in those with CCI 3-4, and 7.1%, SD 1.3 in those with CCI ≥5, p=0.182).

### The factors associated with poor glycaemic control in logistic regression analysis

The unadjusted and adjusted ORs of factors associated with poor glycaemic control are presented in Table 2. In the adjusted model, the odds of having poor glycaemic control were twice as high in participants with polypharmacy compared to those without polypharmacy (adjusted OR 2.18, 95% CI 1.38 – 3.42). Compared to robust participants, the adjusted ORs for poor glycaemic control were 1.75 (95% CI 1.10 – 2.76) in pre-frail participants, and 2.03 (95% CI 1.11 – 3.72) in frail participants. Compared to participants with a diabetes duration from 1 – 5 years, the odds of poor glycaemic control were 2.46 (95% CI 0.97 – 6.21) in those that had diabetes for less than 1 year, 1.68 (95% CI 1.06 – 2.66) in those that had diabetes from 6 to 10 years, and 2.23 (95% CI 1.39 – 3.56) in those that had diabetes for more than 10 years.

**Table 2.**
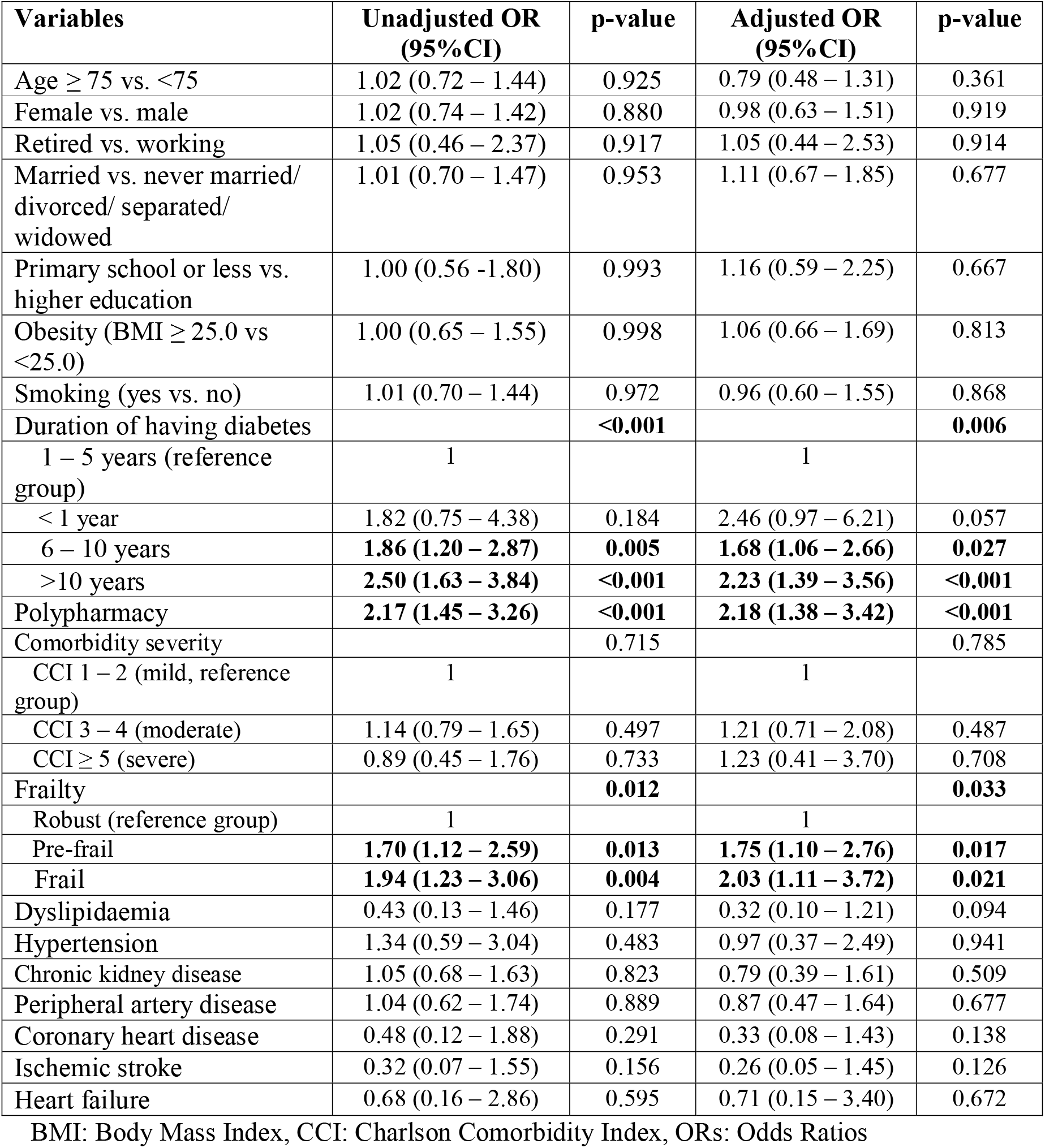
Factors associated with poor glycaemic control.

## Discussion

In our study, nearly half of the study participants had poor glycaemic control. Previous studies in Vietnam and other countries in the Southeast Asia also reported a very high prevalence of poor glycaemic control among adults with T2DM. In a study of 189 patients with T2DM at an urban hospital in Hanoi, Vietnam from 2018 to 2019, 70% of the study participants (mean age 62.4 years, SD 7.6) had uncontrolled glycaemic levels (defined as HbA1c level ≥ 6.5%).^11^ In a study of 557 T2DM patients recruited from seven clinics in Malaysia (mean age 56.0, SD 9.1), 77% had poor glycaemic control (HbA1c ≥ 6.5%), with a mean HbA1c level for all participants of 8.0%, SD 2.0).^17^ Another study of 385 patients with T2DM in 1 general and 17 community hospitals in Thailand (mean age 59.8 years, SD 9.1) found that 80% of the participants had poor glycaemic control (HbA1c ≥ 7%).^18^ In a recent review of the prevalence and management of hypertension in adults with diabetes in Southeast Asian countries, out of the 51 studies that reported the percentages of participants achieving target glucose control, 46 studies found that more than 50% of the participants had poor glucose control.^19^

The two most common comorbidities in this study were dyslipidaemia and hypertension, which was similar to another study that reported hypertension as the most common comorbidity in older adults with diabetes presenting at primary care in Australia.^20^ Of note, more than one third of our study participants were classified as having moderate or severe comorbidity. We did not find any significant difference in the prevalence of the cardiovascular risk factors and comorbidities between participants with good and poor glycaemic control in our study. There are growing number of studies discussing the impact of comorbidities on overall management and progression of T2DM.^21^ The increasing awareness of comorbidities can help assess the risk and benefits of intensifying, maintaining or de-intensifying glucose control treatments in older people with diabetes.^22^ Older people with diabetes face a wide range of medical conditions, functional and cognitive impairment.^23^ Additional variation in psychosocial environment and access to resources further complicates diabetes management for older adults.^24^ Recognizing this variability is crucial for providing effective and person-centred care for them.

Our study found that duration of diabetes, frailty, and polypharmacy were significantly associated with poor glycaemic control among the participants. The mean HbA1c level was highest in participants with diabetes for less than 1 year. This was different when compared to a study in Malaysia^17^ which reported that, for each 1-year increase in duration of diabetes, there was a 5% reduction in the odds of achieving target glycaemic control. This could be due to the mean age of study population in Malaysia being much younger (59.8 years) than in our study (71.9 years). Our finding may suggest further support is needed for older patients with newly diagnosed T2DM. Due to age-related changes and impact, such as cognitive impairment, retirement, and mobility issues, older adults may have difficulties navigating through a diagnosis of diabetes and its treatment when compared to younger population.

The increased odds of having HbA1c ≥ 7.0% among prefrail and frail participants could reflect the cautious approach of the physicians when treating older patients with frailty. In fact, the American Diabetes Association Standards of Care in Diabetes recommended a more relaxed target (HbA1c < 8.0%) for older adults with T2DM and frailty.^8^ The impact of frailty on glycaemic control in our study participants highlights the value of frailty assessment in diabetes. Strain et al. proposed that frailty, rather than age, can influence the prognosis for older adults with diabetes and hence, should be an important factor for setting treatment goals and individualizing care.^25^

The high prevalence of polypharmacy across all participants in our study, regardless of whether they had good or poor glycaemic control highlights further opportunities for improvement. Lipska et al. concluded from four large randomized clinical trials that more aggressive attempts to manage glycaemic levels using multiple pharmacological agents are often associated with reduced benefits and increased potential harms in older adult adults with type 2 diabetes.^13^ Complex medical regimens have been reported to negatively impact patient medication adherence.^26^ The International Geriatric Diabetes Society recently recommended that healthcare providers should consider simpler and safer medication regimens for older adults with diabetes.^6^ Therefore, exploring strategies to simplify treatment plans and reduce polypharmacy could be helpful in improving glycaemic control in older adults with diabetes and multiple comorbidities.

### Strengths and limitations

The strengths of this study lie in its high-quality, multi-centre data and clinical relevance, making it valuable for understanding the impact of common geriatric syndromes, like frailty and polypharmacy, on glycaemic control. However, due to the cross-sectional nature of the study, we could not clarify whether frailty or polypharmacy precedes poor glycaemic control or vice versa. Future longitudinal studies could provide helpful information on this topic. Although this study reported on the association of polypharmacy and glycaemic control, information on type of medicines and adherence were not collected. Future studies could also explore the use of oral antihyperglycaemic agents and insulin together with levels of treatment adherence and its association with glycaemic control. The study participants were recruited from two major hospitals in Ho Chi Minh City, an urban city in Vietnam. Therefore, the findings should be interpreted within this context and may not be generalisable to other regions, such as rural areas.

## Conclusion

There was a high prevalence of poor glycaemic control and high burden of comorbidity, frailty, and polypharmacy among the participants. Diabetes duration, frailty, and polypharmacy were factors significantly associated with glycaemic control among the participants of this study. The study findings highlight the importance of considering geriatric related issues as part of the comprehensive diabetes management and underscore the need for further research on optimizing polypharmacy, frailty and developing personalized treatment plans in managing long-term diabetes.

## Data Availability

All data produced in the present study are available upon reasonable request to the authors

## References

1. Sinclair A, Saeedi P, Kaundal A, Karuranga S, Malanda B, Williams R. Diabetes and global ageing among 65–99-year-old adults: Findings from the International Diabetes Federation Diabetes Atlas, 9th edition. Diabetes Research and Clinical Practice. 2020/04/01/ 2020;162:108078. doi:10.1016/j.diabres.2020.108078

2. The L. Diabetes: a defining disease of the 21st century. The Lancet. 2023;401(10394):2087. doi:10.1016/S0140-6736(23)01296-5

3. Sun H, Saeedi P, Karuranga S, et al. IDF Diabetes Atlas: Global, regional and country-level diabetes prevalence estimates for 2021 and projections for 2045. Diabetes Research and Clinical Practice. 2022/01/01/ 2022;183:109119. doi:10.1016/j.diabres.2021.109119

4. The Lancet Healthy L. Care for ageing populations globally. The Lancet Healthy Longevity. 2021;2(4):e180. doi:10.1016/S2666-7568(21)00064-7

5. Aschner P, Gagliardino JJ, Ilkova H, et al. Persistent poor glycaemic control in individuals with type 2 diabetes in developing countries: 12 years of real-world evidence of the International Diabetes Management Practices Study (IDMPS). Diabetologia. 2020/04/01 2020;63(4):711–721. doi:10.1007/s00125-019-05078-3

6. Munshi M, Kahkoska AR, Neumiller JJ, et al. Realigning diabetes regimens in older adults: a 4S Pathway to guide simplification and deprescribing strategies. The Lancet Diabetes & Endocrinology. 2025;

7. Bellary S, Kyrou I, Brown JE, Bailey CJ. Type 2 diabetes mellitus in older adults: clinical considerations and management. Nature Reviews Endocrinology. 2021/09/01 2021;17(9):534–548. doi:10.1038/s41574-021-00512-2

8. Committee ADAPP. 13. Older Adults: Standards of Care in Diabetes—2025. Diabetes Care. 2024;48(Supplement_1):S266–S282. doi:10.2337/dc25-S013

9. Ngoc NB, Lin ZL, Ahmed W. Diabetes: What Challenges Lie Ahead for Vietnam? Ann Glob Health. Jan 2 2020;86(1):1. doi:10.5334/aogh.2526

10. Vuong TB, Tran TM, Tran NQ. High prevalence of prediabetes and type 2 diabetes, and identification of associated factors, in high-risk adults in Vietnam: A cross-sectional study. Diabetes Epidemiology and Management. 2025/01/01/ 2025;17:100239. doi:10.1016/j.deman.2024.100239

11. Thuy LQ, Nam HTP, An TTH, et al. Factors Associated with Glycaemic Control among Diabetic Patients Managed at an Urban Hospital in Hanoi, Vietnam. BioMed Research International. 2021;2021(1):8886904. doi:10.1155/2021/8886904

12. Tao J, Gao L, Liu Q, et al. Factors contributing to glycemic control in diabetes mellitus patients complying with home quarantine during the coronavirus disease 2019 (COVID-19) epidemic. Diabetes Research and Clinical Practice. 2020/12/01/ 2020;170:108514. doi:10.1016/j.diabres.2020.108514

13. Lipska KJ, Krumholz H, Soones T, Lee SJ. Polypharmacy in the Aging Patient: A Review of Glycemic Control in Older Adults With Type 2 Diabetes. JAMA. 2016;315(10):1034–1045. doi:10.1001/jama.2016.0299

14. Charlson M, Szatrowski TP, Peterson J, Gold J. Validation of a combined comorbidity index. Journal of clinical epidemiology. 1994;47(11):1245–1251.

15. Fried LP, Tangen CM, Walston J, et al. Frailty in older adults: evidence for a phenotype. The journals of gerontology series a: biological sciences and medical sciences. 2001;56(3):M146–M157.

16. Gnjidic D, Hilmer SN, Blyth FM, et al. Polypharmacy cutoff and outcomes: five or more medicines were used to identify community-dwelling older men at risk of different adverse outcomes. Journal of Clinical Epidemiology. 2012/09/01/ 2012;65(9):989–995. doi:10.1016/j.jclinepi.2012.02.018

17. Ahmad NS, Islahudin F, Paraidathathu T. Factors associated with good glycemic control among patients with type 2 diabetes mellitus. J Diabetes Investig. Sep 2014;5(5):563–9. doi:10.1111/jdi.12175

18. Phuwilert P, Khiewkhern S, Phajan T, et al. Factors Affecting Glycemic Control in Patients with Type 2 Diabetes in Kalasin Province, Thailand: An Analytical Cross-Sectional Study. Healthcare. 2024;12(19):1916.

19. Wong WJ, Nguyen TV, Ahmad F, et al. Hypertension in Adults With Diabetes in Southeast Asia: A Systematic Review. J Clin Hypertens (Greenwich). Jan 2025;27(1):e14936. doi:10.1111/jch.14936

20. Wong WJ, Nguyen T, Fortin M, Harrison C. Prevalence and patterns of comorbidities in older people with type 2 diabetes in Australian primary care settings. Australasian Journal on Ageing. 2024;43(2):306–313. doi:10.1111/ajag.13282

21. Hussain S, Chowdhury TA. The Impact of Comorbidities on the Pharmacological Management of Type 2 Diabetes Mellitus. Drugs. 2019/02/01 2019;79(3):231–242. doi:10.1007/s40265-019-1061-4

22. Huang ES. Management of diabetes mellitus in older people with comorbidities. Bmj. 2016;353

23. Munshi MN, Meneilly GS, Rodríguez-Mañas L, et al. Diabetes in ageing: pathways for developing the evidence base for clinical guidance. The Lancet Diabetes & Endocrinology. 2020;8(10):855–867.

24. Leung E, Wongrakpanich S, Munshi MN. Diabetes Management in the Elderly. Diabetes Spectr. Aug 2018;31(3):245–253. doi:10.2337/ds18-0033

25. Strain WD, Down S, Brown P, Puttanna A, Sinclair A. Diabetes and Frailty: An Expert Consensus Statement on the Management of Older Adults with Type 2 Diabetes. Diabetes Ther. May 2021;12(5):1227–1247. doi:10.1007/s13300-021-01035-9

26. Kassaw AT, Sendekie AK, Minyihun A, Gebresillassie BM. Medication regimen complexity and its impact on medication adherence in patients with multimorbidity at a comprehensive specialized hospital in Ethiopia. Original Research. Frontiers in Medicine. 2024-May-27 2024;Volume 11 - 2024 doi:10.3389/fmed.2024.1369569

